# Impact of Modifiable Risk Factors and APOE on Neuropsychiatric Symptoms in Alzheimer’s Disease

**DOI:** 10.64898/2026.06.04.26353599

**Authors:** Hasib Mia, Pamela Del Rosario, Ajneesh Kumar, Nicholas R. Ray, Jiji T. Kurup, Masood Manoochehri, Colin Stein, Alyssa N. De Vito, Brenna Cholerton, Robert A. Sweet, Michael L. Cuccaro, Gary W. Beecham, Edward D. Huey, Christiane Reitz

## Abstract

**BACKGROUND:** Neuropsychiatric symptoms (NPS) are prevalent and debilitating in Alzheimer’s disease (AD). Existing pharmacologic treatments are often ineffective and associated with serious adverse events. Identifying modifiable risk factors (MRFs) is critical for prevention and treatment.

**METHODS:** Capitalizing on data from 14,497 individuals with AD from the National Alzheimer’s Coordinating Center (NACC) database, we examined longitudinal associations between modifiable risk factors, *APOE* genotype and NPI-Q-assessed NPS using Cox proportional hazards models adjusted for demographics.

**RESULTS:** Diabetes, alcohol consumption, smoking, and TBI were associated with an increased risk of specific NPS in AD. *APOEε4* carrier status was linked to multiple NPS, showing a dose-response relationship. Education, LDL-C, and corrective lenses were protective; hypertension showed no associations.

**CONCLUSION:** These findings strongly suggest that individual MRFs are associated with specific NPS in line with a complex etiology underlying these symptoms. Early detection and management of vascular, lifestyle and sensory factors could reduce NPS.

## BACKGROUND

Alzheimer’s disease (AD) is a progressive neurodegenerative disorder characterized by cognitive deterioration, psychiatric symptoms, and changes in behavior. [1] It remains one of the most pressing public health challenges globally, with nearly seven million people affected within the United States and nearly 55 million more worldwide. [2,3] This burden is projected to increase rapidly in the coming decades, posing considerable strain for both the health care systems as well as families and caregivers. [4]

The clinical hallmark features of AD are memory loss and cognitive impairment. However, neuropsychiatric symptoms (NPS) are nearly as pervasive, affecting approximately 85% of individuals with the disease. [1] NPS in AD include agitation, depression, anxiety, hallucinations, delusions, apathy, and disturbances in nighttime behavior. While NPS often appear early and can intensify as the disease progresses, there is temporal variability in these symptoms. Presence of NPS is linked to a cascade of adverse outcomes in the AD course including faster cognitive and functional decline, increased mortality, earlier institutionalization, and significantly greater caregiver burden and distress. [5] Despite their high prevalence and detrimental clinical impact, effective treatments for NPS remain limited with most available pharmacologic options offering only modest benefits and carrying substantial risks, including heightened mortality.[6] Consequently, identification of risk factors for these symptoms, in particular modifiable risk factors (MRFs), is critical to reduce both the prevalence and course of these highly debilitating symptoms in individuals with cognitive impairment. [6]

The Lancet Commission on Dementia Prevention, Intervention, and Care recently published a set of fourteen MRFs that account for a sizable portion of global dementia cases. [4] These factors include limited educational attainment, high LDL cholesterol, hearing loss, hypertension, obesity, smoking, depression, physical inactivity, diabetes, low social contact, excessive alcohol consumption, traumatic brain injury (TBI), air pollution and vision loss. [4] The Lancet Commission estimated that addressing these risk factors during the life course would prevent or delay approximately 45%. (i.e., nearly half) of dementia cases, strongly arguing that policies targeting these factors at an early stage and continuing throughout the life course would have a significant impact on dementia prevalence. Whether this would, however, also reduce risk of specific NPS in AD remains unclear. In the present study we capitalized on data from over 14,697 subjects from the National Alzheimer’s Disease Coordinating Center (NACC), the premier centralized repository for clinical, biomarker, genetic, and neuropathologic data in the United States, to assess whether these modifiable dementia risk factors are associated with risk of individual NPS or specific clusters of NPS in AD using longitudinal time-to-event analyses.

## METHODS

### Participants

Data were obtained from the NACC Uniform Data Set (UDS). The NACC was established in 1999 as the central repository for the Alzheimer’s Disease Research Centers (ADRCs) by the National Institute on Aging in the United States. Inclusion criteria for the current analyses included (1) a diagnosis of AD, (2) Neuropsychiatric Inventory Questionnaire (NPI-Q) data available and (3) data on the Lancet Commission dementia risk factors available yielding 14,697 individuals in the current analyses.

All participants completed informed consent procedures at their individual institutions. The NACC repository was approved by the University of Washington Institutional Review Board.

### AD diagnosis

AD diagnosis was made by site clinician(s) according to criteria specified by NACC (naccdata.org): (1) the participant had cognitive or behavioral symptoms that interfere with independent activities of daily living, represent a decline from previous levels of function, are not explained by a delirium or major psychiatric disorder, and include cognitive impairment noted by the clinician during history-taking and objective cognitive assessment; and (2) the participant had impairment in one or more domains (acquiring or remembering new information, reasoning/handling complex tasks/judgment, visuospatial abilities, language, or changes in personality, behavior, or comportment).[7] The Clinical Dementia Rating (CDR^®^) Dementia Staging Instrument was used to determine stage/severity of dementia. [8] The CDR^®^ is a semi-structured interview and is administered by a clinician or other trained health professional to both the participant and co-participant. It assesses a range of cognitive symptoms and impacts of cognitive changes on activities of daily living (memory, orientation, judgement/problem solving, community affairs, home/hobbies, and personal care), resulting in a global rating score (0 = none, 0.5 = questionable, 1 = mild, 2 = moderate, 3 = severe). The score is based on decline from previous levels of functioning due to cognitive loss rather than changes in physical function or other factors.

### Neuropsychiatric phenotypes

Neuropsychiatric symptoms at each visit were assessed using the NPI-Q, a measure of the severity of 12 NPS common in AD adapted from the full NPI. [9,10] The NPI-Q is one of the most used measures of NPS in AD and has been cross-validated with the NPI and shown good test-retest reliability. [10] In NACC, the NPI-Q is administered by a clinician or other trained health professional to a co-participant (generally a spouse or adult child) and assesses whether the participant has experienced a range of psychiatric symptoms (delusions, hallucinations, agitation/aggression, depression, anxiety, elation/euphoria, apathy, irritability, motor disturbance, nighttime behaviors, and appetite) during the month prior to the study visit. Each NPS is coded as absent or present by a rater and if present is given a severity rating from 1 (mild) to 3 (severe). Co-participants are advised only to endorse a symptom (yes/no) if it occurred following the initial onset of cognitive symptoms.

### Risk Factors

Out of the 14 risk factors validated by the Lancet Commission, eleven were available in the NACC dataset.[4] Risk factors were defined using multiple variables in the dataset and were only coded as present if identified at or prior to the visit when AD was first diagnosed. Each risk factor was treated as a binary variable (1 = present, 0 = absent), unless otherwise stated.

Education was analyzed as a continuous variable representing the total years of formal schooling. Education levels were assigned as follows: 12 = high school or GED, 16 = bachelor’s degree, 18 = master’s degree, 20 = doctoral degree. Hearing impairment was defined using the HEARING and HEARAID variables. Since the HEARING variable was reverse coded in the dataset where 0 indicated normal hearing and 1 indicated hearing impairment, it was recoded so that so that 1 represented impairment and 0 represented no impairment. Similarly, participants who reported using a hearing aid (HEARAID = 1) were coded as having hearing impairment. Participants with other or missing values were excluded. High LDL cholesterol was considered present if participants reported a diagnosis of hypercholesteremia or indicated current use of any lipid-lowering medications. If any of the following variables: HYPCHOL, HYPERCHO and NACCLIPL had a value of 1, high LDL cholesterol was coded as present. Depression was coded as present if either DEP2YRS or DEPOTHR = 1 to capture lifetime incidence of depression. Both variables had to be absent (value = 0) for the risk factor to be coded as absent. Traumatic Brain Injury (TBI) was identified using several variables. Participants were classified as having a history of TBI if any of the following variables, TBI, TBIBRIEF, TRAUMABRF, TBIEXTEN, or TRAUMAEXT, had values of 1 or 2, indicating a current or past brain injury. Diabetes was coded as present if any of the following variables indicated Type I or Type diabetes: DIABETES, DIABTYPE (values of 1 or 2), DIABET (values of 1 or 2) NACCBMD.

Participants coded as 3 (other diabetes types) were excluded from diabetes-related analyses due to diagnostic ambiguity. Smoking status was defined based on calculated pack-year exposure. Participants were coded as having a history of smoking through smoking intensity and duration which were quantified using two variables: SMOKYRS, which reports the total number of years the participant smoked, and PACKSPER, which reports the average number of cigarette packs smoked per day using a categorical scale. These two variables were used to create a derived continuous variable, PACKYRS, calculated by multiplying the number of smoking years by the midpoint of the categorical pack per day range. Participants missing data for either SMOKYRS or PACKSPER were coded as missing for PACKYRS. Smoking was treated as a continuous variable and those with incomplete data on PACKYRS were coded as missing for this risk factor. Hypertension was determined using both diagnostic and medication data. It was coded as present if participants had a diagnosis indicated by HXHYPER or HYPERTEN (values of 1 or 2), or if they were taking any hypertensive medications, including NACCAHTN, NACCHTNC, NACCACEI, NACCAAAS, NACCBETA, NACCCCBS, MACCDIUR, NACCVASD, or NACCANGI. Obesity was defined using the NACCBMI variable. Participants were coded as obese if (BMI) ≥ 30.0 kg/m^2^ recorded at or before the AD diagnosis visit. Excessive alcohol use was considered present if the ALCOHOL variable was coded as 1 (recent/active) or 2 (remote/inactive), with a value of 0 indicating no excessive use. Vision impairment was identified using VISION and VISWCORR. Since VISION was reverse coded, it was recoded so that 1 indicated impairment and 0 indicated no impairment. Impairment was coded present if either variable was 1, indicating that vision was not functionally normal without corrective lenses.

### Statistical Analyses

We conducted Cox proportional hazards models to examine associations between the eleven risk factors and the onset of individual neuropsychiatric symptoms, as measured by the Neuropsychiatric Inventory Questionnaire (NPI-Q), among individuals with AD. Baseline was defined as the time of AD diagnosis, and individuals were followed from baseline until the first occurrence of each neuropsychiatric symptom, last follow-up, or death, whichever occurred first. The time-to-event variable was defined as the time between baseline and symptom onset or censoring. For each NPI-Q item (e.g., delusions, hallucinations, agitation, etc.), separate Cox regression models were fit with the presence of the specific symptom as the event of interest. Participants without the symptom by the end of follow-up were censored. Models were adjusted for age, race, education, and sex. All analyses were conducted using R version 4.4.2. (R Foundation for Statistical Computing, Vienna, Austria). Statistical significance was defined as a two-sided p-value <0.05. As the analyses are hypothesis generating, they were not corrected for multiple testing.

## RESULTS

### Sample Characteristics

The final analytic sample included 14,697 participants who received a clinical diagnosis of AD and met inclusion criteria. The mean age at AD diagnosis was 78.3 years (SD = 9.90) and 54.7% (*n* = 8,036) of the sample were females. The racial composition was as follows: 12,256 (83.40 %) non-Hispanic White, 1,676 (11.40%) Black or African American, 330 (2.25) Asian, 104 (0.71%) American Indian or Alaska Native, and 16 (0.11%) Native Hawaiian or Other Pacific Islander and 315 (2.13%) missing responses. There were 1,180 (8.03%) Hispanic individuals in this sample. Participants had a mean education of 15.5 years (SD = 7.18). 36.00% (*n* = 5,294) were carriers of at least one *APOEε4* allele.

The most frequently observed MRF in the total sample was vision impairment, reported as 80.02%. Hearing loss was reported by 29.48% of the sample. A sizable portion of the sample also had a history of elevated LDL (60.09%) and hypertension (64.11%), two common vascular RFs associated with AD. A documented history of depression was reported in 41.33%. Obesity (18.58%), diabetes (13.81%) and TBI (12.40%) were present to a lesser extent as was alcohol consumption (5.48%).

The most prevalent NPS in this sample of individuals with AD and information on MRFs were irritability (61.84%), apathy (61.79%), anxiety (61.71%) and depression (59.20%). Agitation (55.82%), appetite changes (50.58%), and nighttime behavioral disturbances (54.34%) were only slightly less frequent, indicating at least half of the cohort experiencing at least one of these NPS. Less prevalent symptoms, but still notable were disinhibition (37.60%), motor disturbances (37.14%), delusions (28.51%) and hallucinations (18.39%). Elation (10.89%) was the least commonly observed symptom among participants.

### Cox Proportional Hazards Analyses

Figure 1 and Supplementary Table 1 show the results from the Cox proportional hazards analyses relating each individual MRF with individual NPS outcomes adjusted for sex, age, and ethnic group and education.

**Figure 1.**
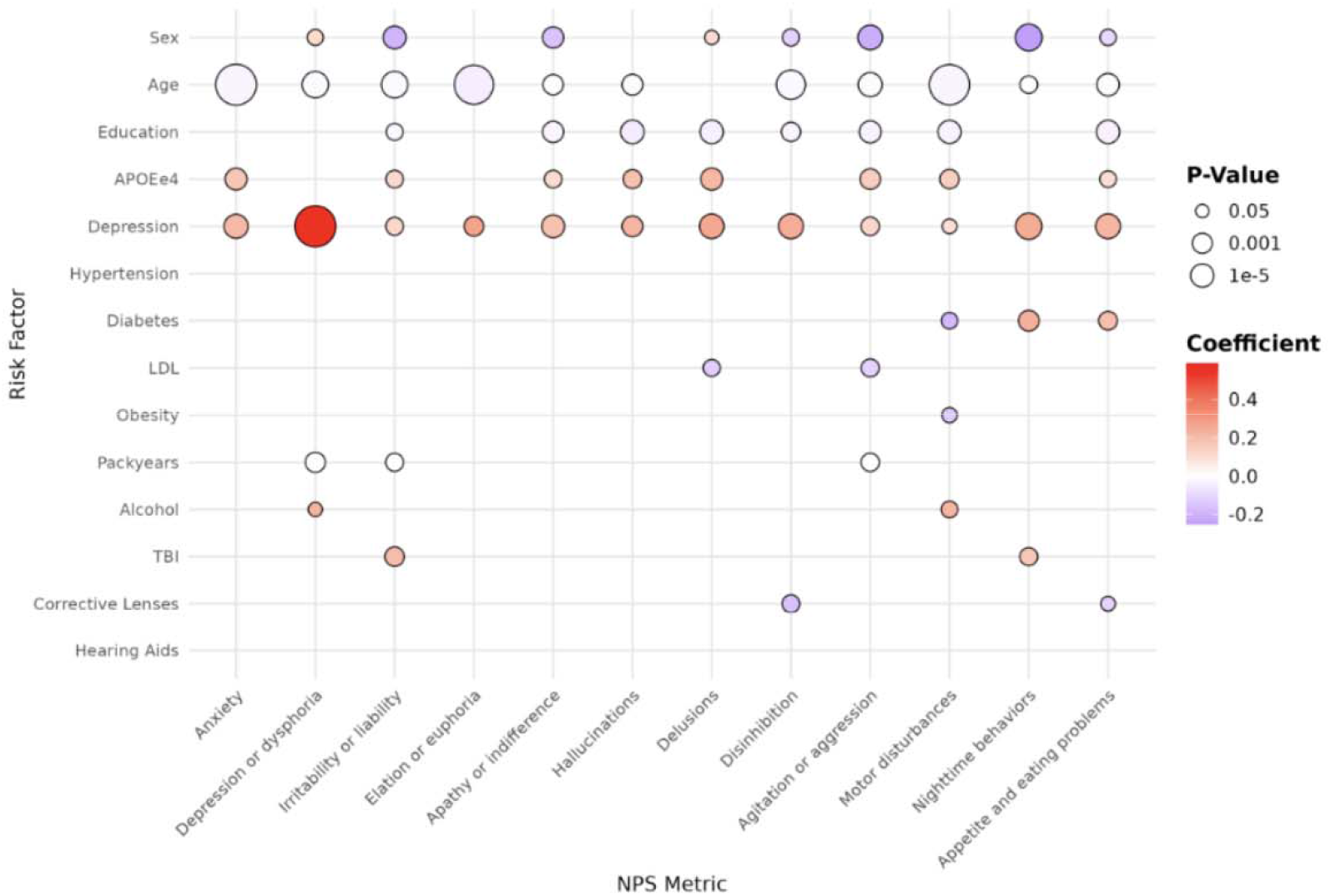
Results from Cox Proportional Hazards Models Examining Associations Between Modifiable Risk Factors and Onset of Neuropsychiatric Symptoms in Individuals with Alzheimer’s Disease. Each circle represents the estimated regression coefficient for each RF-NPS pair. The warmer colors indicate higher risk, while cooler colors indicate protectiveness. Circle size corresponds to statistical significance, with larger circles indicating smaller p-values.

Diabetes was associated with a higher risk of appetite changes and nighttime behaviors. Each additional pack-year of smoking was associated with a statistically significant, modest increase in the risk of agitation, depression, and irritability, consistent with the small effect size expected for a per-year exposure measure. Excessive alcohol use predicted a greater hazard for depression, and motor disturbances. A history of traumatic brain injury was associated with elevated risk for irritability, and nighttime behaviors. A prior history of depression consistently predicted an increased risk across nearly all neuropsychiatric symptom outcomes, including agitation, anxiety, apathy, appetite changes, delusions, depression, disinhibition, elation, hallucinations, irritability, motor disturbances, and nighttime behaviors. Carrying one *APOEε4* allele was significantly associated with increased risk of appetite changes, delusions, hallucinations, irritability, motor disturbances, agitation, anxiety, and apathy. Being homozygous for *APOEε4* was associated with delusions, depression, hallucinations, motor disturbances, agitation, anxiety, and apathy. *APOEε4* homozygotes consistently exhibited larger hazard ratios than heterozygotes, indicating a dose-response relationship between *APOEε4* allele burden and NPS risk.

Several risk factors demonstrated protective effects. Higher educational attainment was consistently associated with a reduced likelihood of agitation, apathy, appetite changes, delusions, disinhibition, hallucinations, irritability, and motor disturbances. Elevated LDL cholesterol appeared protective against agitation and delusions. Both obesity and diabetes were linked to a decreased risk of motor disturbances. Maintained sensory function, indicated by the use of corrective lenses, was associated with a reduced risk of appetite changes and disinhibition.

## DISCUSSION

Leveraging longitudinal data from nearly 15,000 individuals from NACC, this study examined whether MRFs identified by the Lancet Commission on Dementia are associated not only with cognitive decline but also with the onset of specific NPS in individuals with AD. [4] We identified distinct patterns of risk and protection across a range of NPS and MRFs, underscoring the complex mechanistic etiology underlying these debilitating symptoms in AD.

Presence of diabetes, smoking, excessive alcohol consumption, TBI, at least one *APOEε4* allele and depression were associated with an increased risk of specific NPS. Several of the assessed MRFs including diabetes, smoking, alcohol consumption and TBI may contribute to individual NPS in AD via neuroinflammation caused by increased inflammatory cytokine signaling, affecting synaptic plasticity and neuronal function in brain regions involved in behavior and sleep. [11–13] In vivo PET imaging studies of the translocator protein (TSPO), a marker of microglial activation, suggests that neuroinflammation in brain is associated with NPS even after accounting for potential confounders including stage and severity of cognitive impairment or amyloid and tau pathology. [14,15] This is consistent with prior evidence demonstrating that inflammatory profiles are associated with risk and severity of psychiatric disorders including depression and post-traumatic stress disorder. [16,17] Elevated levels of IL-6 and TNF-α, commonly observed in diabetes and chronic smoking, have been associated with impaired neuronal signaling. [13,18] Increased expression of proinflammatory cytokines and inflammatory response proteins may also lead to insulin resistance in the hypothalamus and suprachiasmatic nucleus (SCN), affecting appetite and sleep duration. [19,20]

Chronic smoking may also contribute to NPS in AD through its cumulative effects on brain chemistry and neurovascular health. Nicotine consumption stimulates the release of serotonin, dopamine, and norepinephrine and chronic exposure may lead to receptor desensitization and neurotransmitter depletion, increasing susceptibility to irritability and mood changes.[21] Additionally, smoking is associated with microstructural changes of cerebral white matter cortical areas involved in executive function, memory, and emotional regulation. [22,23] Prolonged excessive alcohol consumption can also disrupt the balance between inhibitory and excitatory neurotransmitters, resulting in emotional instability, irritability, and depression. [24] In addition, alcohol-related neurotoxicity in the frontal cortex and cerebellum can impair motor coordination and behavioral regulation. [25] Finally, focal, or diffuse damage to neurotransmitter-producing and modulating brain regions caused by TBI or cerebrovascular disease can result in NPS including irritability and nighttime behavior through disruption of white matter tracts, diffuse axonal damage, HPA dysregulation, or injury to hypothalamic or brainstem sleep control systems. [26]

While we cannot exclude the possibility that the increased risks of NPS detected by the NPI-Q associated with pre-existing depression are due to shared etiologic mechanisms, depression can disrupt circadian rhythms and REM sleep causing sleep dysfunction that can in turn result in agitation, irritability, nighttime behavioral disturbances, and hallucinations. [27] In addition, depressive symptoms can lead to cognitive bias and misinterpretation of reality and increase risk of suspiciousness, delusions, and misperceptions, and can lead to withdrawal with reduced engagement in social and cognitive activity potentially contributing to apathy. [28,29]

Our findings on associations of the *APOEε4* allele with NPS are in line with a recently reported GWAS for NPI-Q domains in 12,800 participants from NACC that reported genome-wide significant signals for agitation, anxiety, apathy, delusions, and hallucinations that were driven by the *APOE*ε4 allele. [30] Notably, we consistently observed dose-response effects with progressively higher risk of individual NPS among homozygous *APOE*ε4 allele carriers compared to heterozygous carriers. The *APOE* gene encodes apolipoprotein E involved in lipid transport and metabolism and promotes clearance of amyloid-beta (Aβ) from the brain. Besides its mechanistic role in the pathogenesis of cognitive impairment in AD, Aβ pathology has also been associated with an increased risk of NPS. [31] In addition, *APOEε4* can amplify microglial reactivity promoting a pro-inflammatory state, reducing microglia activity to phagocytose Aβ, and disrupting lipid metabolism. [32] As described above, this can exacerbate neuroinflammation and lead to neuronal damage and synaptic dysfunction. [33]

Several MRFs were inversely associated with specific NPS. Consistent with the cognitive reserve hypothesis, but regarding psychiatric symptoms, higher educational attainment was associated with a reduced risk for most NPS. [34] Education may promote more enhanced neural plasticity and executive function, supporting emotion regulation, and impulse control. [35] Neuroimaging studies also suggest that individuals with higher cognitive reserve demonstrate more robust functional connectivity in key regions such as the prefrontal cortex and hippocampus, potentially buffering against symptoms such as agitation, disinhibition, or depression. [36,37] Our findings also indicate that maintaining functional sensory abilities— specifically using corrective lenses—is associated with a reduced risk of select NPS including appetite changes and disinhibition. Preserved sensory input may further support network integrity (e.g., default mode, salience and frontoparietal networks), reducing or preventing disruption in these networks that can lead to affective and behavioral symptoms. [38,39] Wearing corrective lenses may also prevent visual distortions that can promote hyperexcitability and disorganized processing in sensory cortices, increasing susceptibility to spontaneous perceptual activity interpreted as hallucinations. As the effects of sensory impairment itself were not directly assessed, we cannot directly conclude from our study that sensory impairment adversely impacts NPS risk. However, our results strongly suggest that retaining functional sensory capacity may confer protective effects against some neuropsychiatric complications even if impairment is not causally harmful, a critical observation informing preventative public health strategies.

Plasma LDL levels were inversely associated with agitation and delusions. There is evidence showing that these factors have a U-shaped relationship with AD prevalence and incidence over the life course, with presence in midlife being associated with an increased risk of late-life AD, while presence in late life shows an inverse association. [40,41] The same is likely true for clinical subfeatures of AD including specific NPS.

Finally, while diabetes increased risk for some NPS it also appeared to exert a protective effect on motor disturbances. The same association was observed for obesity. Older adults with diabetes and metabolic syndrome commonly receive intensive cardiometabolic management, including insulin and metformin medications. Rodent models show that metformin may exert neuroprotective effects through AMP□activated protein kinase (AMPK) activation and improved neurovascular integrity, potentially limiting secondary neuroinflammatory damage in specific brain regions. [42] Insulin also supports motor function indirectly by enhancing skeletal muscle glucose uptake for energy and by modulating autonomic motor circuits through hypothalamic pathways. [43] Individuals with obesity have also higher circulating leptin levels which can promote neuronal survival and synaptogenesis potentially preserving motor pathways. [44]

We did not observe an association of hypertension with risk of any NPS in our study although evidence from epidemiologic, genetic, and experimental studies suggests that hypertension is a MRF for cognitive impairment in AD. [45] Hypertension-induced cerebrovascular damage disproportionately affects white matter tracts and cortical-subcortical connectivity and has less impact on monoaminergic nuclei and their projections, the fronto-limbic network, and synaptic/neurotransmitter regulation. [46]

Furthermore, there were no association of hearing aids with risk of any NPS in our study. While hearing aid use partially restores auditory perception, the spiral ganglion neurons (SGN) often suffer irreversible damage limiting the extent to which peripheral amplification can normalize auditory input. [47] Hearing loss is also associated with structural (insula and amygdala) and functional (anterior cingulate salience network) changes in brain regions involved in emotion and behavior. [48] Therefore, even when hearing aids are utilized, the downstream effects of cortical-limbic circuits may be dysregulated.

This study has several notable strengths. It capitalizes on a large, well-characterized longitudinal cohort of 14,697 individuals from NACC with longitudinal standardized data, allowing time-to-event modeling with high statistical power. Limitations are that two Lancet Commission AD RFs, physical activity, and air pollution, were not collected in NACC and could therefore not be assessed in this study, and that NACC includes subjects engaged with specialized research centers, with participants being more educated, healthier, and less ethnically diverse compared to the general population. This can affect generalizability of our results to the broader community.

Our findings have considerable clinical and public health implications. Although NPS can be one of the earliest signs of neurodegenerative disease, they are often underdiagnosed due to inaccuracies in self-reporting and lack of awareness in clinical settings, which can result in devastating outcomes for both patients and their caregivers. [49] When NPS are appropriately diagnosed, they are often treated off label by psychotropic medications that have limited efficacy and significant adverse effects including increased mortality. [50] Our findings strongly suggest that early detection and management of vascular and lifestyle factors including diabetes, smoking, alcohol consumption, and TBI, as well as maintaining sensory function and promoting higher educational attainment would reduce the risk of NPS in individuals with AD. Studies are needed that further clarify the specific molecular mechanism through which these factors increase the risk of specific NPS.

**Table 1.** *Sample Characteristics (N = 14,697)*. Data are presented as n (%), and (mean ± SD) unless otherwise noted.

| Characteristics | Frequency |
| --- | --- |
| Total Number of Participants | 14,697 |
| Age at Diagnosis (mean ± SD) | 78.3 ± 9.90 |
| **Gender** |  |
| Female | 8,036 (54.70%) |
| Male | 6,661 (45.30%) |
| **Race** |  |
| White | 12,256 (83.40 %) |
| Black or African American | 1,676 (11.40%) |
| American Indian or Alaska Native | 104 (0.71%) |
| Native Hawaiian or Other Pacific Islander | 16 (0.11%) |
| Asian | 330 (2.25) |
| N/A | 315 (2.13%) |
| **Years of Education (mean ± SD)** | 15.50 ± 7.18 |
| **APOEE4 Genotype** |  |
| -/- | 5,639 (38.40%) |
| e4/- | 5,294 (36.00%) |
| e4/e4 | 1,456 (9.91%) |
| N/A | 2,308 (15.70%) |
| **Risk Factors** |  |
| Hearing | 4333 (29.48%) |
| LDL | 8,832 (60.09%) |
| Depression | 6,075 (41.33%) |
| TBI | 1,822 (12.40%) |
| Diabetes | 2,030 (13.81%) |
| Hypertension | 9423 (64.11%) |
| Obesity | 2,731 (18.58%) |
| Excessive Alcohol Consumption | 805 (5.48%) |
| Vision | 11761 (80.02%) |
| Pack Years (mean ± SD) | 15.5 ± 28.7 |
| **Neuropsychiatric Symptoms** |  |
| Delusions | 4,191 (28.51%) |
| Hallucinations | 2,703 (18.39%) |
| Agitation | 8,204 (55.82%) |
| Depression | 8,701 (59.20%) |
| Anxiety | 9,069 (61.71%) |
| Elation | 1,601 (10.89%) |
| Apathy | 9,082 (61.79%) |
| Disinhibition | 5,526 (37.60%) |
| Irritability | 9,089 (61.84%) |
| Motor Disturbances | 5,459 (37.14%) |
| Nighttime Behaviors | 7,986 (54.34%) |
| Appetite Changes | 7,434 (50.58%) |

## Data Availability

All data utilized in this study were obtained from the National Alzheimer's Coordinating Center (NACC). The NACC Uniform Data Set (UDS) is available to researchers upon submission of a formal data request and approval from their committee. Data can be requested directly through their website at https://naccdata.org/.

https://www.naccdata.org/data-request-process/

