## Supplemental Table 1 for "Impact of Modifiable Risk Factors and APOE on Neuropsychiatric Symptoms in Alzheimer’s Disease"

|  |  |  | **HR** | **SE** | **P Value** |
| --- | --- | --- | --- | --- | --- |
| **ANXIETY** | **no anxiety (n (%))** | **anxiety (n (%))** |  |  |  |
| Education | 15.9 (2.8) | 16 (2.7) | 0.992 | 0.008 | 3.18E-01 |
| e4/- | 1545 (34.1%) | 1453 (39.4%) | 1.180 | 0.050 | 3.56E-04 |
| e4/e4 | 388 (8.6%) | 453 (12.3%) | 1.210 | 0.070 | 3.83E-03 |
| Depression | 1413 (31.1%) | 1413 (38.3%) | 1.245 | 0.042 | 1.71E-07 |
| Hypertension | 3033 (66.9%) | 2192 (59.4%) | 0.935 | 0.045 | 1.36E-01 |
| Diabetes | 640 (14.1%) | 440 (11.9%) | 0.946 | 0.067 | 4.08E-01 |
| LDL | 2717 (59.9%) | 2159 (58.5%) | 0.998 | 0.041 | 9.55E-01 |
| Obesity | 850 (18.7%) | 586 (15.9%) | 0.885 | 0.057 | 3.13E-02 |
| Pack Years | 13.5 (27.5) | 11.9 (24.9) | 0.999 | 0.001 | 4.54E-01 |
| Alcohol Consumption | 232 (5.1%) | 195 (5.3%) | 1.076 | 0.094 | 4.33E-01 |
| Traumatic Brain Injury | 501 (11%) | 446 (12.1%) | 1.077 | 0.065 | 2.54E-01 |
| Corrective Lenses | 3613 (79.6%) | 2919 (79.1%) | 0.952 | 0.054 | 3.62E-01 |
| Hearing Aids | 1388 (30.6%) | 972 (26.3%) | 0.987 | 0.051 | 1.09E+00 |
| **DEPRESSION** | **no depression (n (%))** | **depression (n (%))** |  |  |  |
| Education | 16 (2.7) | 15.7 (2.8) | 0.983 | 0.008 | 3.86E-02 |
| e4/- | 1855 (36.9% | 1092 (36.6%) | 0.940 | 0.050 | 2.50E-01 |
| e4/e4 | 454 (9.0%) | 368 (12.3%) | 1.160 | 0.070 | 4.11E-02 |
| Depression | 3345 (37.4%) | 1170 (45.6%) | 1.810 | 0.047 | 3.16E-36 |
| Hypertension | 3179 (63.2%) | 1806 (60.5%) | 1.038 | 0.050 | 4.61E-01 |
| Diabetes | 4825 (54.0%) | 1358 (52.9%) | 1.021 | 0.046 | 6.51E-01 |
| LDL | 5375 (60.2%) | 1466 (57.1%) | 0.996 | 0.045 | 9.28E-01 |
| Obesity | 1607 (18.0%) | 409 (15.9%) | 1.067 | 0.060 | 2.81E-01 |
| Pack Years | 12.5 (25.6) | 12.4 (24.3) | 1.003 | 0.001 | 1.54E-03 |
| Alcohol Consumption | 467 (5.2%) | 148 (5.8%) | 1.306 | 0.102 | 9.22E-03 |
| Traumatic Brain Injury | 551 (10.9%) | 382 (12.8%) | 1.123 | 0.072 | 1.09E-01 |
| Corrective Lenses | 3971 (78.9%) | 2389 (80%) | 1.032 | 0.062 | 6.12E-01 |
| Hearing Aids | 1448 (28.8%) | 815 (27.3%) | 1.107 | 0.055 | 6.56E-02 |
| **IRRATIBILITY** | **no irritability (n (%))** | **irritability (n (%))** |  |  |  |
| Education | 15.9 (2.7) | 15.9 (2.8) | 0.981 | 0.008 | 9.88E-03 |
| e4/- | 1620 (35.2%) | 1383 (39.2%) | 1.120 | 0.050 | 1.37E-02 |
| e4/e4 | 446 (9.7%) | 432 (12.2%) | 1.120 | 0.070 | 1.08E-01 |
| Depression | 1603 (34.9%) | 1358 (38.5%) | 1.145 | 0.043 | 1.43E-03 |
| Hypertension | 2831 (61.6%) | 2100 (59.5%) | 1.002 | 0.047 | 9.73E-01 |
| Diabetes | 548 (11.9%) | 444 (12.6%) | 1.024 | 0.065 | 7.12E-01 |
| LDL | 2647 (57.6%) | 2029 (57.5%) | 1.016 | 0.042 | 6.96E-01 |
| Obesity | 746 (16.2%) | 578 (16.4%) | 0.973 | 0.058 | 6.39E-01 |
| Pack Years | 11.5 (25.8) | 11.6 (23.8) | 1.002 | 0.001 | 4.14E-02 |
| Alcohol Consumption | 199 (4.3%) | 177 (5.0%) | 1.236 | 0.096 | 2.81E-02 |
| Traumatic Brain Injury | 513 (11.2%) | 455 (12.9%) | 1.220 | 0.064 | 2.01E-03 |
| Corrective Lenses | 3641 (79.2%) | 2780 (78.8%) | 0.951 | 0.055 | 3.64E-01 |
| Hearing Aids | 1295 (28.2%) | 932 (26.4%) | 1.028 | 0.052 | 5.93E-01 |
| **ELATION** | **no elation (n (%))** | **elation (n (%))** |  |  |  |
| Education | 15.9 (2.8) | 15.8 (2.7) | 0.980 | 0.015 | 1.71E-01 |
| e4/- | 4209 (36.8%) | 382 (40.1%) | 1.080 | 0.090 | 3.98E-01 |
| e4/e4 | 1195 (10.4%) | 111 (11.7%) | 0.870 | 0.140 | 2.85E-01 |
| Depression | 4538 (39.7%) | 453 (47.6%) | 1.337 | 0.080 | 2.89E-04 |
| Hypertension | 7215 (63.1%) | 540 (56.7%) | 0.916 | 0.089 | 1.09E+00 |
| Diabetes | 1535 (13.4%) | 104 (10.9%) | 0.828 | 0.135 | 1.63E-01 |
| LDL | 6767 (59.2%) | 557 (58.5%) | 1.046 | 0.081 | 5.74E-01 |
| Obesity | 2011 (17.6%) | 166 (17.4%) | 0.864 | 0.109 | 1.79E-01 |
| Pack Years | 12.7 (25.9) | 11.5 (22.5) | 0.999 | 0.002 | 5.32E-01 |
| Alcohol Consumption | 606 (5.3%) | 70 (7.4%) | 1.499 | 0.149 | 6.56E-03 |
| Traumatic Brain Injury | 1382 (12.1%) | 125 (13.1%) | 1.023 | 0.123 | 1.30E+00 |
| Corrective Lenses | 9085 (79.4%) | 733 (77%) | 0.923 | 0.104 | 1.13E+00 |
| Hearing Aids | 3247 (28.4%) | 216 (22.7%) | 1.115 | 0.104 | 1.37E+00 |
| **APATHY** | **no apathy (n (%))** | **apathy (n (%))** |  |  |  |
| Education | 16 (2.8) | 15.9 (2.8) | 0.973 | 0.007 | 1.12E-04 |
| e4/- | 1611 (36.0%) | 1552 (38.6%) | 1.100 | 0.040 | 3.93E-02 |
| e4/e4 | 401 (9.0%) | 480 (12.0%) | 1.180 | 0.070 | 9.99E-03 |
| Depression | 1436 (32.1%) | 1548 (38.5%) | 1.221 | 0.040 | 6.55E-07 |
| Hypertension | 2785 (62.2%) | 2502 (62.3%) | 0.957 | 0.067 | 5.11E-01 |
| Diabetes | 572 (12.8%) | 484 (12.1%) | 1.031 | 0.062 | 6.21E-01 |
| LDL | 2683 (60.0%) | 2313 (57.6%) | 0.953 | 0.039 | 2.18E-01 |
| Obesity | 815 (18.2%) | 655 (16.3%) | 0.994 | 0.054 | 9.15E-01 |
| Pack Years | 12 (24.2) | 12.2 (26.3) | 1.001 | 0.001 | 1.26E-01 |
| Alcohol Consumption | 213 (4.8%) | 199 (5.0%) | 0.893 | 0.095 | 2.35E-01 |
| Traumatic Brain Injury | 523 (11.7%) | 507 (12.6%) | 0.894 | 0.078 | 1.48E-01 |
| Corrective Lenses | 3604 (80.6%) | 3178 (79.1%) | 1.100 | 0.040 | 3.93E-02 |
| Hearing Aids | 1298 (29%) | 1074 (26.7%) | 1.180 | 0.070 | 9.99E-03 |
| **HALLUCINATION** | **no hallucination (n (%))** | **hallucination (n (%))** |  |  |  |
| Education | 16 (2.7) | 15.6 (2.8) | 0.949 | 0.010 | 5.82E-07 |
| e4/- | 3792 (36.7%) | 773 (39.9%) | 1.180 | 0.070 | 1.16E-02 |
| e4/e4 | 1044 (10.1%) | 261 (13.5%) | 1.270 | 0.090 | 1.05E-02 |
| Depression | 4038 (39.1%) | 900 (46.5%) | 1.293 | 0.058 | 8.60E-06 |
| Hypertension | 6470 (62.6%) | 1166 (60.2%) | 0.946 | 0.064 | 3.87E-01 |
| Diabetes | 1316 (12.7%) | 271 (14.0%) | 1.073 | 0.088 | 4.22E-01 |
| LDL | 6146 (59.5%) | 1112 (57.4%) | 0.918 | 0.057 | 1.37E-01 |
| Obesity | 1823 (17.6%) | 320 (16.5%) | 0.892 | 0.078 | 1.42E-01 |
| Pack Years | 12.7 (25.9) | 11.5 (23.8) | 0.999 | 0.001 | 5.70E-01 |
| Alcohol Consumption | 568 (5.5%) | 101 (5.2%) | 0.878 | 0.136 | 3.42E-01 |
| Traumatic Brain Injury | 1303 (12.6%) | 228 (11.8%) | 1.028 | 0.091 | 1.23E+00 |
| Corrective Lenses | 8240 (79.7%) | 1523 (78.6%) | 0.941 | 0.078 | 1.10E+00 |
| Hearing Aids | 2968 (28.7%) | 475 (24.5%) | 0.964 | 0.074 | 6.14E-01 |
| **DELUSION** | **no delusion (n (%))** | **delusion (n (%))** |  |  |  |
| Education | 16 (2.7) | 15.7 (2.8) | 0.960 | 0.009 | 5.57E-06 |
| e4/- | 3213 (36.0%) | 1027 (40.0%) | 1.210 | 0.060 | 8.30E-04 |
| e4/e4 | 865 (9.7%) | 353 (13.7%) | 1.350 | 0.080 | 1.61E-04 |
| Depression | 5375 (60.0%) | 1466 (57.0%) | 1.312 | 0.050 | 4.23E-08 |
| Hypertension | 1124 (13.0%) | 319 (12.0%) | 0.936 | 0.055 | 2.24E-01 |
| Diabetes | 3345 (37.0%) | 1170 (46.0%) | 1.042 | 0.077 | 5.89E-01 |
| LDL | 3823 (43.0%) | 1068 (42.0%) | 0.897 | 0.049 | 2.66E-02 |
| Obesity | 4825 (54.0%) | 1358 (53.0%) | 0.880 | 0.068 | 5.89E-02 |
| Pack Years | 1607 (18.0) | 409 (16.0) | 1.001 | 0.001 | 1.78E-01 |
| Alcohol Consumption | 8099 (91.0%) | 2303 (90.0%) | 1.094 | 0.110 | 4.14E-01 |
| Traumatic Brain Injury | 1115 (12.5%) | 316 (12.3%) | 1.060 | 0.077 | 4.50E-01 |
| Corrective Lenses | 7125 (79.8%) | 2059 (80.2%) | 1.050 | 0.068 | 4.76E-01 |
| Hearing Aids | 2579 (28.9%) | 686 (26.7%) | 0.990 | 0.061 | 8.72E-01 |
| **DISINHIBITION** | **no disinhibition (n (%))** | **disinhibition (n (%))** |  |  |  |
| Education | 15.9 (2.7) | 15.8 (2.8) | 0.976 | 0.008 | 3.10E-03 |
| e4/- | 2810 (36.3%) | 1152 (39.1%) | 1.030 | 0.050 | 6.22E-01 |
| e4/e4 | 768 (9.9%) | 369 (12.5%) | 0.940 | 0.080 | 4.26E-01 |
| Depression | 2868 (37.0%) | 1280 (43.4%) | 1.265 | 0.046 | 3.08E-07 |
| Hypertension | 4874 (62.9%) | 1752 (59.5%) | 0.927 | 0.050 | 1.32E-01 |
| Diabetes | 996 (12.9%) | 363 (12.3%) | 1.043 | 0.072 | 5.56E-01 |
| LDL | 4579 (59.1%) | 1719 (58.3%) | 0.928 | 0.045 | 1.01E-01 |
| Obesity | 1307 (16.9%) | 514 (17.4%) | 1.017 | 0.061 | 7.83E-01 |
| Pack Years | 12.1 (25.3) | 12.5 (25.4) | 1.002 | 0.001 | 1.06E-02 |
| Alcohol Consumption | 362 (4.7%) | 172 (5.8%) | 1.216 | 0.100 | 5.13E-02 |
| Traumatic Brain Injury | 906 (11.7%) | 394 (13.4%) | 1.114 | 0.069 | 1.20E-01 |
| Corrective Lenses | 6201 (80.1%) | 2289 (77.7%) | 0.856 | 0.059 | 8.71E-03 |
| Hearing Aids | 2185 (28.2%) | 793 (26.9%) | 1.045 | 0.057 | 4.37E-01 |
| **AGITATION** | **no agitation (n (%))** | **agitation (n (%))** |  |  |  |
| Education | 16 (2.7) | 15.9 (2.8) | 0.973 | 0.007 | 1.49E-04 |
| e4/- | 3213 (36.0%) | 1027 (40.0%) | 1.150 | 0.050 | 2.88E-03 |
| e4/e4 | 865 (9.7%) | 353 (13.7%) | 1.220 | 0.070 | 2.76E-03 |
| Depression | 1863 (35.2%) | 1568 (40.1%) | 1.150 | 0.040 | 5.31E-04 |
| Hypertension | 3320 (62.7%) | 2355 (60.2%) | 0.959 | 0.044 | 3.45E-01 |
| Diabetes | 653 (12.3%) | 471 (12.0%) | 0.981 | 0.063 | 7.65E-01 |
| LDL | 3184 (60.1%) | 2246 (57.4%) | 0.896 | 0.039 | 5.41E-03 |
| Obesity | 924 (17.5%) | 632 (16.2%) | 0.946 | 0.055 | 3.07E-01 |
| Pack Years | 11.4 (24.1) | 12.9 (26) | 1.003 | 0.001 | 1.64E-03 |
| Alcohol Consumption | 244 (4.6 %) | 202 (5.2 %) | 1.243 | 0.089 | 1.50E-02 |
| Traumatic Brain Injury | 630 (11.9%) | 495 (12.7%) | 1.064 | 0.062 | 3.14E-01 |
| Corrective Lenses | 4233 (79.9%) | 3107 (79.4%) | 0.958 | 0.054 | 4.28E-01 |
| Hearing Aids | 1508 (28.5%) | 1047 (26.8%) | 1.019 | 0.049 | 7.04E-01 |
| **MOTOR DISTURBANCE** | **no motor disturbance (n (%))** | **motor disturbance (n (%))** |  |  |  |
| Education | 16 (2.8) | 15.8 (2.8) | 0.964 | 0.008 | 2.62E-06 |
| e4/- | 2737 (35.5%) | 1330 (40.2%) | 1.140 | 0.050 | 8.61E-03 |
| e4/e4 | 700 (9.1%) | 469 (14.2%) | 1.200 | 0.070 | 7.01E-03 |
| Depression | 2917 (37.9%) | 1423 (43%) | 1.121 | 0.044 | 8.99E-03 |
| Hypertension | 4968 (64.5%) | 1954 (59%) | 0.928 | 0.048 | 1.19E-01 |
| Diabetes | 1036 (13.5%) | 381 (11.5%) | 0.877 | 0.073 | 7.14E-02 |
| LDL | 4631 (60.1%) | 1913 (57.8%) | 0.937 | 0.043 | 1.34E-01 |
| Obesity | 1403 (18.2%) | 546 (16.5%) | 0.845 | 0.059 | 4.49E-03 |
| Pack Years | 12.8 (25.7) | 11.7 (24.6) | 1.001 | 0.001 | 4.87E-01 |
| Alcohol Consumption | 387 (5%) | 203 (6.1%) | 1.329 | 0.088 | 1.23E-03 |
| Traumatic Brain Injury | 959 (12.5%) | 398 (12%) | 0.890 | 0.070 | 9.75E-02 |
| Corrective Lenses | 6197 (80.5%) | 2604 (78.6%) | 0.972 | 0.059 | 6.26E-01 |
| Hearing Aids | 2278 (29.6%) | 856 (25.8%) | 1.042 | 0.055 | 4.55E-01 |
| **NIGHTIME BEHAVIORS** | **no nighttime behaviors (n (%))** | **nighttime behaviors (n (%))** |  |  |  |
| Education | 15.9 (2.8) | 15.9 (2.8) | 0.987 | 0.007 | 7.55E-02 |
| e4/- | 2066 (36.4%) | 1442 (39.3%) | 0.980 | 0.050 | 6.54E-01 |
| e4/e4 | 588 (10.4%) | 419 (11.4%) | 0.980 | 0.070 | 7.10E-01 |
| Depression | 2011 (35.4%) | 1485 (40.4%) | 1.259 | 0.042 | 3.17E-08 |
| Hypertension | 3462 (61%) | 2266 (61.7%) | 1.078 | 0.045 | 9.61E-02 |
| Diabetes | 659 (11.6%) | 494 (13.4%) | 1.264 | 0.062 | 1.70E-04 |
| LDL | 3348 (59%) | 2153 (58.6%) | 0.966 | 0.041 | 3.97E-01 |
| Obesity | 943 (16.6%) | 624 (17%) | 1.076 | 0.054 | 1.75E-01 |
| Pack Years | 12.3 (26.1) | 11.7 (23.6) | 1.000 | 0.001 | 8.37E-01 |
| Alcohol Consumption | 288 (5.1%) | 213 (5.8%) | 1.176 | 0.088 | 6.56E-02 |
| Traumatic Brain Injury | 617 (10.9%) | 486 (13.2%) | 1.179 | 0.062 | 8.44E-03 |
| Corrective Lenses | 4506 (79.4%) | 2857 (77.8%) | 0.945 | 0.054 | 2.99E-01 |
| Hearing Aids | 1525 (26.9%) | 1012 (27.6%) | 1.005 | 0.051 | 9.19E-01 |
| **APPETITE** | **no appetite (n (%))** | **appetite (n (%))** |  |  |  |
| Education | 16 (2.7) | 15.8 (2.8) | 0.966 | 0.007 | 1.87E-06 |
| e4/- | 2154 (35.7%) | 1468 (39.6%) | 1.110 | 0.050 | 1.92E-02 |
| e4/e4 | 589 (9.7%) | 435 (11.7%) | 1.080 | 0.070 | 2.33E-01 |
| Depression | 2195 (36.3%) | 1510 (40.7%) | 1.244 | 0.041 | 8.91E-08 |
| Hypertension | 3750 (62.1%) | 2269 (61.1%) | 1.029 | 0.045 | 5.23E-01 |
| Diabetes | 744 (12.3%) | 476 (12.8%) | 1.187 | 0.062 | 5.95E-03 |
| LDL | 3552 (58.8%) | 2186 (58.9%) | 1.012 | 0.040 | 7.64E-01 |
| Obesity | 1050 (17.4%) | 646 (17.4%) | 1.038 | 0.054 | 4.90E-01 |
| Pack Years | 12.5 (25.8) | 12.5 (25.9) | 1.001 | 0.001 | 1.32E-01 |
| Alcohol | 707 (11.7%) | 478 (12.9%) | 0.097 | 0.924 | 3.55E-01 |
| Traumatic Brain Injury | 701 (11.6%) | 474 (12.8%) | 1.038 | 0.064 | 5.66E-01 |
| Corrective Lenses | 4825 (79.9%) | 2893 (78%) | 0.897 | 0.054 | 4.22E-02 |
| Hearing Aids | 1761 (29.1%) | 986 (26.6%) | 0.971 | 0.050 | 5.57E-01 |

**Supplementary Table 1***. Results from Cox Proportional Hazards Models Examining Associations Between Modifiable Risk Factors and Onset of Neuropsychiatric Symptoms in Individuals with Alzheimer’s Disease*. All NPS included in the NPI-Q and their associations with Lancet RFs are shown. Education and pack years are reported as mean ± SD; all other variables are presented are as percentages. HR = hazard ratio; larger HR indicates increased risk whereas lower HR indicates a protective effect. SE = standard error. The last column shows p-values, with values at or below 0.05 considered statistically significant.
